# Quantifying SARS-CoV-2 infection risk within the Google/Apple exposure notification framework to inform quarantine recommendations

**DOI:** 10.1101/2020.07.17.20156539

**Authors:** Amanda M. Wilson, Nathan Aviles, James I. Petrie, Paloma I. Beamer, Zsombor Szabo, Michelle Xie, Janet McIllece, Yijie Chen, Young-Jun Son, Sameer Halai, Tina White, Kacey C. Ernst, Joanna Masel

## Abstract

Most Bluetooth-based exposure notification apps use three binary classifications to recommend quarantine following SARS-CoV-2 exposure: a window of infectiousness in the transmitter, ≥15 minutes duration, and Bluetooth attenuation below a threshold. However, Bluetooth attenuation is not a reliable measure of distance, and infection risk is not a binary function of distance, nor duration, nor timing. We model uncertainty in the shape and orientation of an exhaled virus-containing plume and in inhalation parameters, and measure uncertainty in distance as a function of Bluetooth attenuation. We calculate expected dose by combining this with estimated infectiousness based on timing relative to symptom onset. We calibrate an exponential dose-response curve based on infection probabilities of household contacts. The probability of current or future infectiousness, conditioned on how long post-exposure an exposed individual has been symptom-free, decreases during quarantine, with shape determined by incubation periods, proportion of asymptomatic cases, and asymptomatic shedding durations. It can be adjusted for negative test results using Bayes Theorem. We capture a 10-fold range of risk using 6 infectiousness values, 11-fold range using 3 Bluetooth attenuation bins, ∼6-fold range from exposure duration given the 30 minute duration cap imposed by the Google/Apple v1.1, and ∼11-fold between the beginning and end of 14 day quarantine. Public health authorities can either set a threshold on initial infection risk to determine 14-day quarantine onset, or on the conditional probability of current and future infectiousness conditions to determine both quarantine and duration.

## 1. INTRODUCTION

Manual contact tracing followed by quarantine of known contacts is a critical method for containing or mitigating the spread of communicable diseases (Armbruster & Brandeau, 2007). It is, however, extremely resource and time-intensive and relies on case recall of contacts. New technologies can supplement this approach. Manual contact tracing can be effective for COVID-19 (Aleta et al., 2020; Bi et al., 2020; Fetzer & Graeber, 2020; Kendall et al., 2020; Kucharski et al., 2020), however, a significant challenge is the extremely short window of time between an infected individual presenting for testing and the contacts that they infected beginning to shed infectious virus (Ferretti, Wymant, et al., 2020; Kretzschmar et al., 2020). Automatic exposure notification approaches based on Bluetooth proximity have the potential to achieve many of the benefits of contact tracing, while also providing more rapid notification, greater privacy (Fraser et al., 2020; Von Arx et al., 2020), more objective recall of contacts including those whose identity is unknown to the case, and greater scalability (Ferretti, Wymant, et al., 2020; Salathé et al., 2020). The two approaches of contact tracing and exposure notifications are complementary and may directly interact e.g. when those receiving digital exposure notifications are referred to human contact tracers for the information and support needed for quarantine adherence and further investigation (Webster et al., 2020).

Apps have access to data on timing, duration, and Bluetooth attenuation. Determining the threshold for entering quarantine based on probability of infection should yield better results than from combining three binary thresholds for duration, distance, and the infectious period of the transmitter. A threshold for exiting quarantine based on the conditional probability of current or future infectiousness could also be used. Both would help optimize the reduction in disease transmission per day of quarantine recommended.

Here we lay out a framework for doing so using the decentralized protocol of the Google/Apple Exposure Notification (GAEN) Application Programming Interface (API). When a user reports positive infection status, the GAEN framework (Fig. 1) allows apps to assign a “Transmission Risk Level” (version 1) or “infectiousness” (version 2) to each day that they might have been shedding, and to communicate this level to the receiver’s phone via a Temporary Exposure Key (TEK). On the receiver’s device, the GAEN framework records Bluetooth attenuation as a rough estimate of distance, and the duration of exposure.

**Fig 1.**
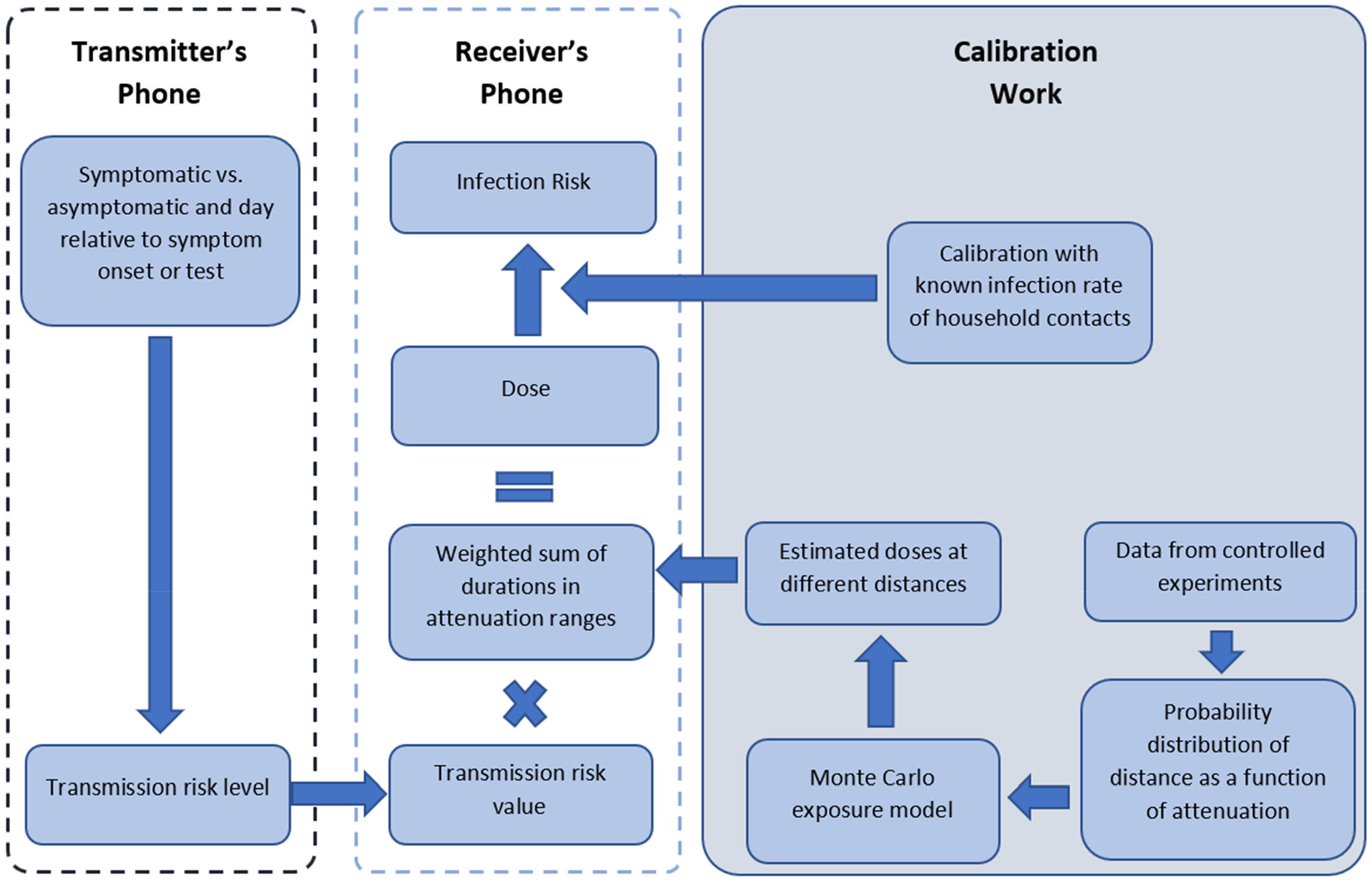
Assessment of the probability of infection following a single exposure. The calibration work is reported in this manuscript, and the procedures on the Transmitter’s and Receiver’s phones are part of the Covid Watch app. The terms “Transmission risk level” and “Transmission risk value” are as used in GAEN v1. In GAEN v2, it is necessary to repurpose “report type” metadata associated with Temporary Exposure Keys and combine it with the two provided levels of “infectiousness” in order to obtain up to 8 levels of infectiousness (Klingbeil, 2020b, 2020a).

The risk of infection depends on viral dose (Haas, Rose, & Gerba, 1999), which in turn depends on the shedding rate of the infected individual, and on the duration and distance of the interaction. As days go by without onset of symptoms, the probability of future infectiousness decreases, because the probability is conditioned on lack of symptoms for an increasing stretch of time. We parameterize calculations of both probabilities using both past literature and new experiments and illustrate what different risk thresholds imply for quarantine recommendations. We are piloting and evaluating the Covid Watch app using portions of this scheme on the campus of the University of Arizona.

## 2. METHODS

The overall approach to calculating infection risk is summarized in Fig. 1. Parameter values and their descriptions and sources are summarized in Supplementary Table I for calculations performed by the app and in Supplementary Table II for parameters we used during calibration.

### 2.1. GAEN Overview

Our experiments were performed in GAEN version 1. However, the method we used is a good simulation of what became standard in the subsequently released version 2. We calculate a weighted sum of durations at different Bluetooth attenuations, using the weights to capture the differences in expected dose (number of inhaled particles over an exposure time). We then multiply by the expected infectiousness of the transmitter, as estimated from the literature in Supplementary Methods Section 2. While this calculation is supported in version 1, access to the necessary data triggers operating system notifications of exposure even when that exposure is minimal, and so is impractical in the field. This calculation became standard in GAEN version 2, in which the dose is referred to in units of “Meaningful Exposure Minutes”. When distance changes over time, accuracy is constrained by the frequency with which GAEN records BlueTooth.

### 2.2. Experiments on Distance-Attenuation Relationship

We measured Bluetooth attenuation for a range of distances, phones, and scenarios of possible signal interference with the potential to affect the attenuation – distance relationship (Supplementary Materials Section 1) (Farrell & Leith, 2020). Using a developer version of the Covid Watch app, we called the API multiple times with different attenuation thresholds in order to achieve resolution of 3dB in the 30dB-99dB range. The API appears to round up durations to 5-minute increments, each with its own attenuation value; we consider each of these to be a datapoint.

Our tests were all short, e.g. a 12-minute test would yield 3 datapoints. This is because the GAEN version 1 framework, under which the experiments were performed, records exposure durations only up to 30 minutes, in order to protect anonymity of COVID-positive patients by limiting the risk that users will be able guess the source of their exposure, while still meeting contact definitions that invoke minimum exposure duration of 15 minutes. This cap has been lifted in GAEN version 2.

There were 7 testers and 14 phones, representing a variety of models, all of iPhones – handset type and orientation can affect signal (Farrell & Leith, 2020). 49 measurements were taken with specific phone orientations, while for the remaining 986 measurements the devices were side-by-side and facing upwards if not otherwise specified by the barrier type (e.g. pocket). 203, 222, 199, 374, 17, 20 measurements were at 0.5m., 1m., 1.5m., 2m., 3m., and 5m., respectively. We also used the 28, 28, 29, 27, and 16 zero-risk barrier measurements at 0.5m., 1m., 1.5m., 2m., and “N/A”, respectively. The phones were stationary during all measurements.

1163 out of the total 1558 datapoints were used for attenuation weight and attenuation threshold setting, with exclusions of data described in Fig. S1. The 1163 non-excluded datapoints are supplied in Supplementary Dataset 1. Of the included 1035 attenuation measures that involved infection risk, 747 did not include a deliberate barrier, while 288 includes barriers such as pockets, backpacks, nearby laptop, and human body. 925 measures were taken inside homes, 49 were taken inside an elevator, and 61 outside.

### 2.3. Setting Attenuation Bin Thresholds and Corresponding Weights

To rebalance the distance measurements to form a pseudo dataset that is more representative of the distribution of barriers and scenarios in the real world, we created a pseudo-dataset with different multiples of the data collected at each of the distances. To inform the desired distribution of distances, we analyzed the time-weighted pairwise distance in traffic flow simulations of a classroom (Jain, Islam, Chowdhury, Chen, & Son, 2021). These indicate a roughly uniform distribution over possible distances, with a reduction in close contact due to attempts to adhere to social distancing rules. Since close contact might be more common in other settings, and distances beyond 5m. can also register Bluetooth signal, we made 5, 5, 6, 3, 132, and 168 copies of the non-zero-risk data at distances of 0.5m., 1m., 1.5m., 2m., 3m., and 5m, respectively, yielding a data ratio of 1015 : 1100 : 1194 : 1122 : 2244 : 3360 (as a rough approximation of a target ratio of 1:1:1:1:2:3) prior to the sampling described below. To this, we added 4 copies of the zero-risk barrier measurements, so that they made up 4.85% of the total pseudo-dataset. Our calibration code holds shedding rate and exposure duration constant at 50 arbitrary units/m^3^ and 30 minutes, in order to isolate the effect of distance on differences in dose between attenuation buckets.

From this pseudo dataset, we first sample a datapoint that falls within the attenuation bin in question. If this is a zero-risk barrier scenario, we assign an infection risk of 0. Otherwise, we record the distance *ρ* in meters. Note that our method is not based on mapping thresholds in distance to thresholds in Bluetooth attenuation, but instead on resampling from the probability distribution of distance as a function of attenuation.

We feed this distance into a microbial exposure model that estimates the airborne spread of viral particles from an emitter’s mouth following a Gaussian plume formation, and their subsequent inhalation by contacts.

#### 2.3.1. Estimation of Exposure Concentrations

It is well-acknowledged that both distance from an infected individual and duration of “close proximity interactions” (Guo et al., 2019) are important parameters in estimating the probability of infection of those exposed (Chu et al., 2020; Rea et al., 2007; Salathé et al., 2010; Setti et al., 2020). However, there is little quantitative information about the relationship between distance and risk of infection. Chu et al. (2020) quantified risk in terms of answers to binary survey questions about whether the respondent came within distance X of an infected person (Chu et al., 2020). They found that the value of the threshold distance X in the survey question predicts the degree to which the answer predicts risk, but this relationship cannot easily be converted into one between actual distance and risk.

For this reason, we instead model the dose inhaled at different distances. Exhaled breath is a likely source of infection (W. Chen, Zhang, Wei, Yen, & Li, 2020; Ma et al., 2020). Accordingly, we model a Gaussian plume (Brusca et al., 2016) of virus-containing aerosols originating from the emitter’s face at (0,0,0). The x axis represents the direction that the transmitter is facing and breathing toward with breath velocity U (m/s). Diffusion causes spread away from y=0 or z=0. The viral concentration is then

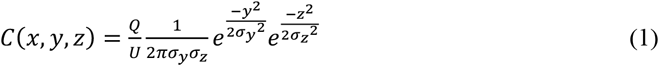

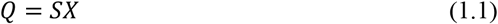

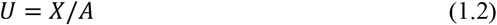

where Q is virus emitted per second and is equal to the product of shedding rate, S, (in arbitrary units proportional to copies/m^3^) and an exhalation rate, X, (taken from measured inhalation rates in m^3^/s), yielding arbitrary units proportional to copies per second being generated (eq 1.1). We sample our exhalation rates from a normal distribution of inhalation rates with a mean and standard deviation of 16.3 and 4.15 m^3^/day, respectively. These were informed by the 16-21 year old range from Table 6-1 in the Exposure Factors Handbook (2011) (U.S. Environmental Protection Agency, 2011). To avoid negative exhalation rates, this distribution was left-truncated at 9 m^3^/day, the smallest fifth percentile of inhalation rates for males and females in age ranges overlapping with the 16-21 year old range (U.S. Environmental Protection Agency, 2011). The velocity of breath U (m/s) was determined by dividing the exhalation rate (m^3^/s) by the cross-sectional area of an open mouth *A* (m^2^), which is the area over which air is assumed to be exhaled at the plume source. The cross-sectional area was informed by a uniform distribution with minimum and maximum cross-sectional areas measured for an open mouth with a “large bite” configuration, ranging from 23 to 59 cm^2^ (Leckie et al., 2000). Note that for a steady-state plume assuming a continuous output of virus, the effects of the exhalation rate (volume of air per second) on amount of virus emitted, and on the velocity with which they disperse, cancel out. For an abrupt exhalation such as a cough, rather than steady state, a higher exhalation rate would affect viral airborne concentration.

For interactions ≤ 1m, we assumed two people interacting are directly in front of each other along the x-axis (φ = π /2, θ=0). For interactions beyond the close range (>1m), we sample θ from a uniform 360 degrees (min=0, max=2π), and the angle between the z axis and the xy-plane, φ, was randomly sampled from a triangular distribution (min=π/4, mode=π/2, max=3π/4). We then convert from spherical units to (x,y,z) to apply Eq. 1. We assumed that scenarios where the person exposed was behind the emitter (x<0) resulted in a zero dose.

To capture the shape of the plume, we use:

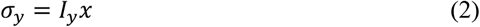

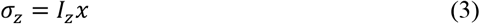

Assuming moderately stable conditions, *I*_*y*_ and *I*_*z*_ were randomly sampled from uniform distributions with minimums and maximums of 0.08-0.25 and 0.03-0.07, respectively (Western Engineering, n.d.).

We note that inhalation and exhalation rates are both likely important to risk. For example, one infected dance instructor spread COVID-19 to 7/26 other instructors at a four hour workshop (Jang, Han, & Rhee, 2020), representing a similar risk as for household contacts, despite the presumption that most were at >2 m. distance for most of this time. Limited air circulation or increased respiratory rates are important factors that cannot be captured in the current GAEN approach, but the four-hour duration of the workshop is captured in GAEN version 2, and when combined with considerable uncertainty in the relationship between Bluetooth attenuation and infection risk per minute, this can appropriately capture the high risk of such a scenario.

While wind velocity and relative humidity are important factors for determining droplet and fine aerosol dispersion and deposition (Feng, Marchal, Sperry, & Yi, 2020; W. Yang, Elankumaran, & Marr, 2011), as is mask usage, these are uncertain factors that are not recorded by the app, especially considering that interactions may occur indoors or outdoors. By not accounting for deposition, and by assuming that masks are either not worn or not worn effectively, we will tend to overestimate dose at greater distances, and in the presence of masks. This will implicitly lower the app-imposed risk tolerance of individuals who comply with public health guidelines that recommend masks and physical distancing, and who might therefore also be more inclined to comply with quarantine recommendations. The 2-meter rule was based on the assumption that most transmission is via droplets (large aerosols) for which deposition occurs over this distance. However, there is increasing evidence for transmission via smaller aerosols (Fennelly, 2020; Jones et al., 2020; Lednicky et al., 2020; Prather, Wang, & Schooley, 2020; Qureshi et al., 2020), supporting our assignment of some risk to greater distances, reflecting short-to medium-distance airborne transmission.

#### 2.3.2. Inhaled Dose per Interaction

An inhaled dose of viral particles due to person-to-person interactions was estimated based on the duration of the interaction (minutes) (T), the concentration of virus in the air at this {x,y,z} coordinate during the interaction (arbitrary units of viral particles/m^3^) C(x,y,z), and inhalation rates (m^3^/minute) (*I*),

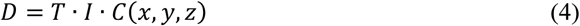

Inhalation rates were randomly sampled from the same distribution as exhalation rates but allowing for a different value per iteration. As with exhalation rates, we left-truncated the distribution to avoid negative inhalation rates and therefore negative doses. Fig. 2 shows the expected dose as a function of distance, with a discontinuity at 1m. arising from our assumption that this distance or below indicates face-to-face interaction.

**Fig 2.**
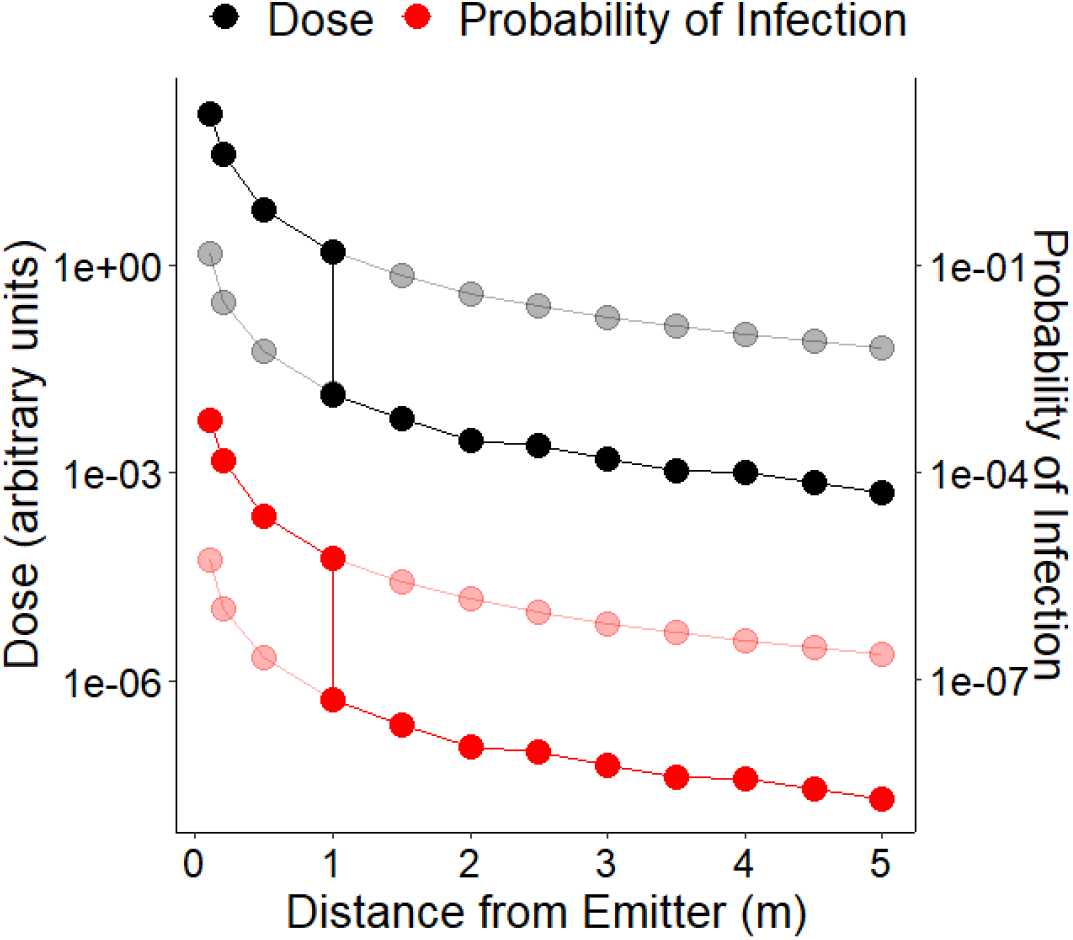
Expected dose and corresponding probability of infection for a 30-minute exposure, estimated using our Monte Carlo procedure as a function of distance from an infected individual. The discontinuity at 1 meter indicates our assumption that this distance threshold indicates face-to-face interaction. Faded points show doses and infection risks that would be estimated if a face-to-face or non-face-to-face interaction assumption were consistent across distances. The bolded points indicate what we assumed in our framework. Note that Bluetooth information likely contains more risk information regarding whether an interaction was face-to-face than it does about risk as a function of the distance at which either a face-to-face or a non-face-to-face interaction takes place. The WHO close contact definition invoking 1 meter also invokes face-to-face interaction (World Health Organization, 2020). The same is true, only with 2 meters, for European guidance (European Centre for Disease Prevention and Control, 2020) The Centers for Disease Control and Prevention (CDC)’s definition departs from this in omitting reference to face-to-face when referring to interactions occurring within 6 feet (Centers for Disease Control and Prevention, 2020).

We use a Monte Carlo approach to sample angle, exhalation rate of the transmitter, cross-section of the transmitter’s open mouth, and inhalation rate of the exposed individual, to obtain a mean dose/time for that attenuation bin. For distances ≤ 1 meter, we assume face to face interactions, consistent with distances measured for “interpersonal” interactions (Zhang et al., 2020). We choose thresholds between attenuation bins, and relative risks for time spent in each bin.

To select the threshold values (a, b) demarcating 3 attenuation bins, we optimized the differences in mean dose between two randomly sampled attenuation measurements. Specifically, we maximized the value of

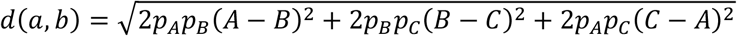

where *A, B* and *C* are the average doses *D* from Eq. 4, averaged across Monte Carlo sampling described above, corresponding to bins [0, a], (a, b], and (b, +), and *p*_*A*_, *p*_*B*_, and *p*_*C*_ are the probabilities that an attenuation will fall within that bin in our pseudo dataset.

We examined multiple local maxima of this distance measure before choosing a partition pair. We also investigated alternative versions of a distance metric and alternative rebalancing schemes, to confirm that this is a relatively robust partition pair.

To relate estimated dose to infection risk, we use an exponential dose-response curve, which is derived from the assumption that each host is susceptible and that each virus has an independent probability of survival and subsequent initialization of infection (Haas et al., 1999). In our case, this probability *k*, multiplied by a constant *C* to convert from arbitrary units to number of virions, sets the parameter *λ = kC* in the equation

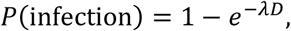

where expected dose *D* comes from a shedding rate multiplied by a weighted sum of time spent within 3 attenuation ranges. An exponential dose-response curve is superior to the approximate beta-Poisson for some other viruses (http://qmrawiki.org/content/recommended-best-fit-parameters, accessed 09/07/2020). These viruses include adenovirus, enterovirus, poliovirus, and SARS-CoV-1.

### 2.4. Calibrating the Dose-Response Curve

Our weighted sum of durations and our estimates of shedding rates *S* in the Results are both in arbitrary units. We therefore fit *λ* to obtain infection probabilities that are compatible with household spread. Asymptomatic infection and low test sensitivity can both deflate estimated household infection risks, while indirect chains of infection via a third household member can inflate them. A meta-analysis by Curmei et al. attempted to correct for these complications and estimated a secondary attack rate of household contacts of 30% (Curmei, Ilyas, Evans, & Steinhardt, 2020). We assumed exposure is equivalent to 8 hours with the maximum shedding rate in the lowest attenuation and calculated *λ* for this dose that would result in a 30% infection risk.

### 2.5. Probability of Current or Future Infectiousness

Our scheme can be used either to 1) set a threshold on the initial probability of infection to trigger 14-day quarantine, or 2) set a threshold for the probability of current or future infectiousness to determine both who should quarantine and for how long. To calculate residual risk of infection as a function of initial risk plus time since exposure, we use the probability distribution of incubation periods from Lauer et al., available at https://iddynamics.jhsph.edu/apps/shiny/activemonitr/ (Lauer et al., 2020). Note that it is possible that incubation periods are even more dispersed than reported here (Wei et al., 2020); this would lengthen quarantine recommendations.

To calculate risk of current or future infectiousness, we assume a fraction of symptomatic vs. asymptomatic cases and take an average of the discount factors applying in each case. Across a population, 20% of infections are estimated to be asymptomatic (Buitrago-Garcia et al., 2020). Younger users are more likely to be asymptomatic (Davies et al., 2020), so the fraction of asymptomatic cases could be personalized on the basis of user age if that information is collected on a voluntary basis. For the symptomatic cases, we discount according to the probability of subsequently developing symptoms, given that symptoms have not appeared yet.

For the asymptomatic cases, we combine the incubation periods from (Lauer et al., 2020) with a distribution of shedding durations from (Long et al., 2020). Long et al. report slightly longer shedding durations for asymptomatic than symptomatic shedding (Long et al., 2020), but other studies for which we were unable to obtain the data, report the opposite, or no difference (Chau et al., 2020; X. Chen et al., 2020; Hu et al., 2020; Xiao et al., 2020; R. Yang, Gui, & Xiong, 2020). Shedding declines in magnitude post symptom onset and is considered by the CDC to have reached negligible levels by 10 days post symptom onset. We assume that asymptomatic shedding begins 3 days before what would have been the day of symptom onset if symptomatic, or else immediately upon infection, whichever occurs later.

Using this assumption, we calculated the probability distribution of the day that shedding ends, given both the distribution of incubation periods and a distribution of shedding durations. For the latter, we combine the asymptomatic and symptomatic shedding durations of (Long et al., 2020) but on the basis of CDC advice for isolation, we truncate the distribution so that all shedding periods longer than 12 days are recorded as exactly 12 days.

Note that low dose exposures, e.g. to asymptomatic individuals, may result in longer incubation periods (Wei et al., 2020), suggesting that low initial risk scores should have longer rather than the shorter quarantines we calculate using this method. We currently ignore this by assuming that risk scores primarily capture uncertainty in the likelihood of infection with a minimal dose, and not variation in the infecting dose once above the minimal. This is supported by genetic evidence in support of an extreme population bottleneck of only 1-8 virions upon transmission, despite using clinical samples data that included presumed high-dose transmission (Lythgoe et al., 2020). We note that lognormal distributions of incubation periods with substantial variance occur even in the absence of variation in dose, due both to variance in within-host replication rate and to the stochastics of establishing infection in the first cells (Ottino-Loffler, Scott, & Strogatz, 2017).

To see how the assumption of negligible variance in infecting dose arises from our model, note that the exponential dose-response curve we use assumes that each virion has an independent probability of initiating infection. Under the resulting Poisson distribution for the number of virions responsible for the initial infection, then even for the 30% infection rate of household contacts, the probability that infection is initiated with two or more virions is only 5%, and with three or more virions is only 0.6%.

However, the higher variance in dose explored in Fig. S2 could make initiation with multiple virions common enough to matter for high infection probabilities. In this case, our simplifying assumption might require overly long quarantines following very high risk exposures. Unless the variance is extreme, it will not significantly distort estimated probabilities among the range of lower risk exposures.

### 2.6. Negative Test Results to Shorten Quarantine

This method can be extended to include the effect of a negative test result on a recommended duration of quarantine. Incorporation of negative test results can help exclude asymptomatic infection and hence allow for earlier release. From Bayes Theorem, and taking the false positive rate as negligible, a negative test result changes the probability of infection from *p* to 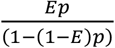, where *E* is the false negative rate. This could be taken as 0.3 (Ai et al., 2020; Y. Yang et al., 2020) or made dependent on the timing of the test relative to exposure (Hellewell et al., 2020; Kucirka, Lauer, Laeyendecker, Boon, & Lessler, 2020).

Kucirka et al. report a false negative rate as a function of the timing of a PCR test relative to symptom onset (Kucirka et al., 2020), but most of the data is post-symptom onset, with only a single patient’s data informing false positive rates prior to symptom onset. The data of (Hellewell et al., 2020) is more suitable for combining with the distribution of incubation periods to calculate the false negative rate as a function of time since exposure, conditional on lack of symptoms to date (Petrie, Nurtay, Ferretti, Fraser, & Masel, 2021). Careful treatment of shared conditionality on symptom onset day enables such calculations even when there are exposures on multiple days (Petrie & Masel, 2021).

### 2.7. Multiple Exposures and Total Risk

Note that strictly speaking when using this latter threshold, our “quarantine” recommendations are, through their treatment of the possibility of undiagnosed asymptomatic infection, a combination of quarantine and isolation. Our scheme, by expressing exposures in terms of probabilities of infection and infectiousness, naturally lends itself to combining risks over multiple exposures. GAEN version 2 sums exposures over 24 periods beginning and ending at midnight UTC). To calculate total risk, we combine the probabilities *p*_*i*_ of each exposure *i*, each discounted as described in the section above, as 1 *-* ∏_*i*_(1 *-p*_*i*_).

Figs. 3 and 4 illustrate scenarios of a single exposure. When there are multiple exposures, quarantine durations are determined with respect to total risk. The risk threshold for initiation and completion of quarantine are the same. In other words, risk is treated in an internally consistent fashion to maximize the benefit from a given number of recommended quarantine days across a population. When fixed quarantine durations are used, exposure must be significant on a single day, from which the 14 days are then calculated, and risks are not integrated across multiple days.

**Fig 3.**
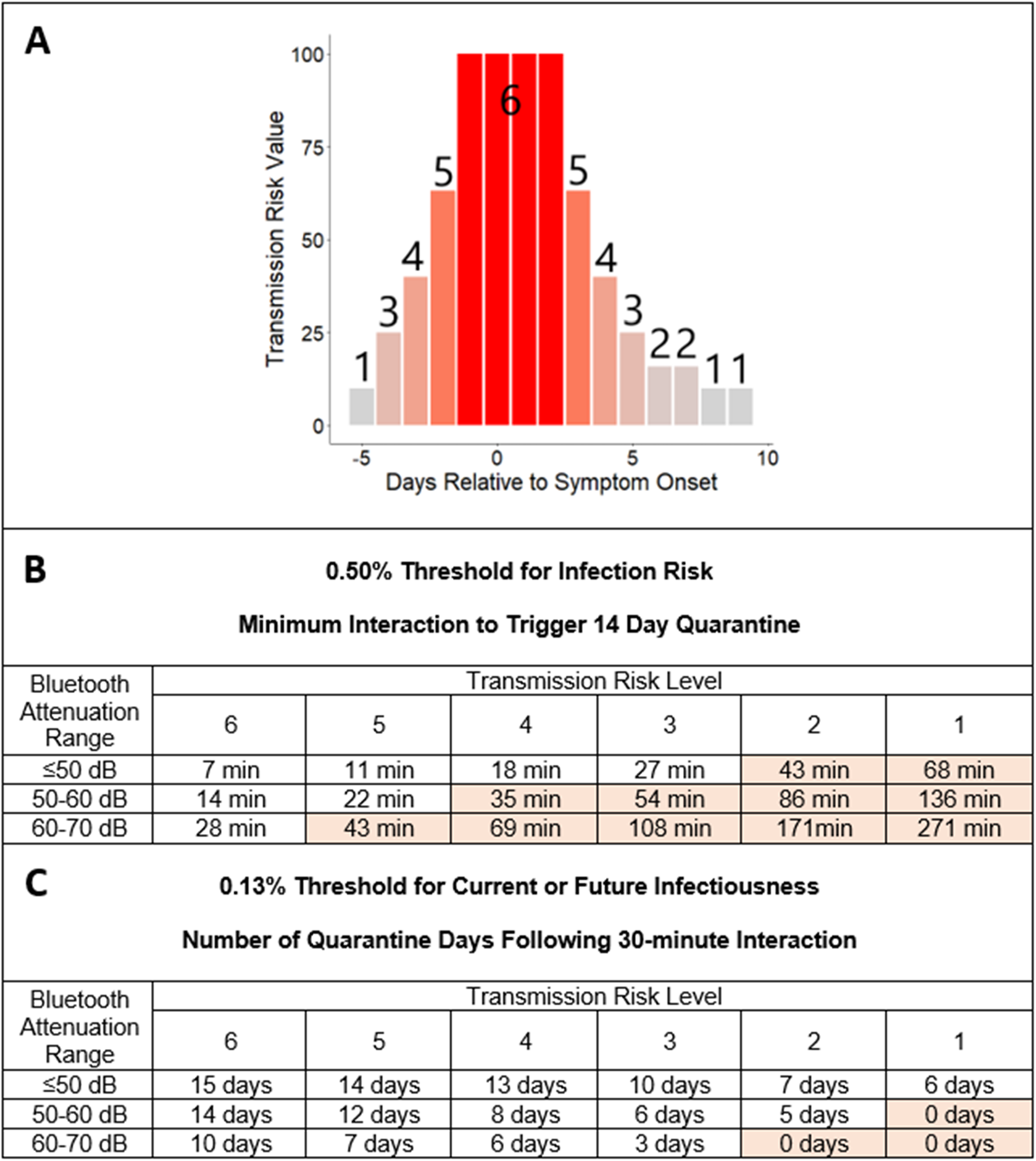
Examples of quarantine recommendations using a threshold for infection risk (B) vs. for current or future infectiousness (C). A) Transmission risk levels 1-6 are used to capture the 10-fold range of relative infectiousness on different days as a function of timing relative to symptom onset. Evidence from both transmission pairs and TCID50 measurements is reviewed in the Supplementary Materials Section 2. B) The minimum length interaction needed to trigger 14-day quarantine is a function both of Bluetooth signal attenuation and of infectiousness. Approaches that neglect the latter correspond to a single row of 15 minutes, and potentially a second row of 30 minutes. Shaded cells indicate that a 30-minute interaction would be insufficient to trigger quarantine, creating issues for GAEN version 1. C) Number of quarantine days recommended following a 30-minute interaction.

**Fig 4.**
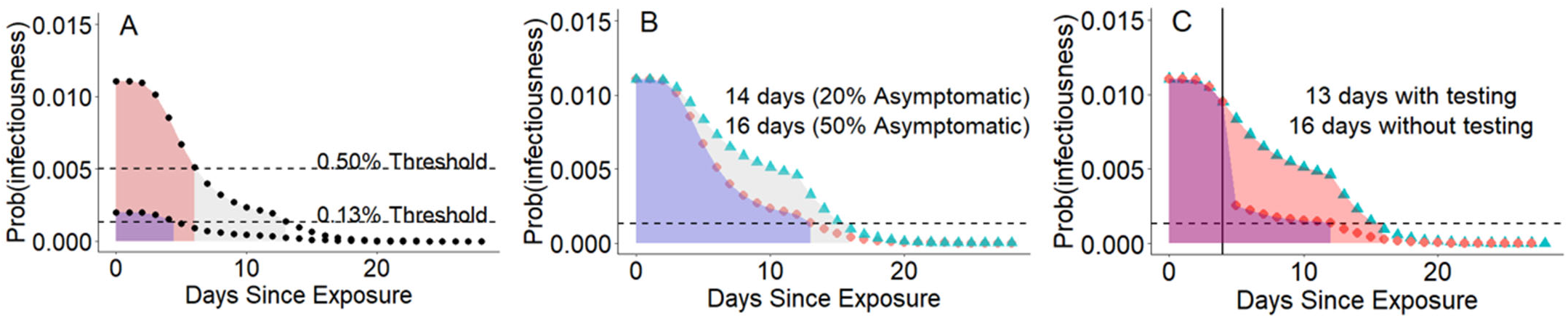
Applying a consistent risk tolerance for current or future infectiousness causes quarantine duration to be a function of initial risk, of the tolerated degree of risk, of the fraction of infections that are assumed to be asymptomatic, and of any negative test results. A) Initial infection risk is 1.10% following 15 minutes of close contact with an individual around the time of symptom onset. With a 20% asymptomatic fraction, a 14-day quarantine is recommended under a 0.13% risk threshold, but only a 7-day quarantine under a 0.5% threshold. Following a lower risk exposure with 0.2% infection risk, quarantine would be 5 days with the stricter threshold, and there would be no quarantine with the less strict. B) Quarantine must be longer to mitigate a high likelihood of asymptomatic infection in the exposed individual. C) A negative test result, shown here as taking place on Day 5, can shorten quarantine, in particular mitigating the risk of asymptomatic infection. We apply Bayes theorem with 70% sensitivity and 100% specificity. Note that widespread availability of testing would allow much stricter risk thresholds to be used. Day 0 is included in the total quarantine times.

## 3. RESULTS

Our Gaussian plume model of microbial exposure produces the relationship between distance and infection risk shown in Fig. 2. Training on both this and our distance-attenuation measurements (as summarized in Methods section 2.2), we chose attenuation bins of ≤ 50dB, 50-60dB, and 60-70 dB, with weights 2, 1, and 0.5, respectively. GAEN version 2 refers to these attenuation bins as Immediate, Near, and Medium. We assign a weight of 0 for >70 dB not because there is no residual infection risk, but because the maximum distance for which BlueTooth signals are still recorded can be highly device-dependent.

Using these weights, we calibrate *λ* = 3.70 × 10^−6^ (see Methods section 2.4) to obtain an infection probability of 0.30 for household contacts. Note that the best way to calibrate both weights and λ would be after the app is rolled out, with manual contact tracers or other opt-in data export compiling exposure characteristics and relating them to the rate of subsequent infection. While Eq. 5 calculates the function of an expectation rather than an expectation of a function, treating variance in dose amounts to using an “effective” value of *λ* (Supplementary Materials Section 1).

Bluetooth attenuation thus only distinguishes a 2-fold difference in dose and hence risk between Immediate and Near range, and only 4-fold between Immediate and Medium range. In contrast, informed both by TCID50 data (Bullard et al., 2020) and by epidemiological evidence (Ferretti, Ledda, et al., 2020), we assign a 10-fold higher risk to exposures to individuals during peak shedding than during the margins of the infectious period (Supplementary Materials Section 2, illustrated in Fig. 3A). The magnitude of shedding (infectiousness) has received less attention than attenuation and exposure duration. It was not widely used by other GAEN apps until version 2 pushed the use of two rather than one levels of infectiousness, determined relative to symptom onset day, and a working group was convened to recommend settings, including input from the current work (Wanger, 2020).

The relatively low predictive power of Bluetooth attenuation gives rise to diagonal patterns in the quarantine recommendations in Fig. 3B. These diagonal patterns mean that quarantine will sometimes be recommended following prolonged exposure to a high shedder, even if the interaction took place at well beyond the estimated 2 m. distance. However, these exposures are not risk-free either, in particular if taking place in an indoor environment, especially in cases with heavy breathing, such as exercise environments (Jang et al., 2020) or choir rehearsals (Hamner et al., 2020), where aerosols may mix throughout the room and also deposit on surfaces. The diagonal pattern reflects the compelling evidence that exposure timing and duration also significantly contribute to infection risk. We therefore sometimes recommend quarantine recommendation even when Bluetooth attenuation, which is a poor proxy for distance, is not low. However, Bluetooth attenuation is nevertheless critical to concluding that an interaction occurred at all.

So far, we have estimated the probability of infection from an exposure. Each day that passes without symptoms provides more information to make infection less likely, and eventually also to increase the probability that shedding from an asymptomatic infection has ended. To calculate the probability of current or future infectiousness on a subsequent day, conditional on no symptoms until that day, we apply a discount factor based both on time elapsed without symptoms and also any negative test results. We multiply the probability of infection from an exposure by this discount factor to determine the remaining risk of infectiousness from a given exposure.

Traditional quarantine guidelines are binary (either 14 days from date of last exposure, or no quarantine required). However, a consistent approach to risk, combined with a desire to impose quarantine days in the most efficient manner possible to combat disease spread, suggests that individuals should quarantine for longer following a higher-risk exposure (Fig. 4A) (although see Methods section 2.5. for a caveat with very high doses). This approach calculates the number of days post-interaction that would be needed to drop below a given threshold of probability of current or future infectiousness. Exposure scenarios of 30 minutes are illustrated in Fig. 3C.

We used a 0.13% threshold in Fig. 3C, because it recommends a 14-day quarantine for 15 minutes in close range with a high shedder. Such an interaction has a 1.10% infection risk, which falls below a 0.13% probability of current or future infectiousness after 14 days of quarantine during which no symptoms appear. Note that this initial infection risk is broadly compatible with the attack rate reported in Taiwan (1.0%, 95% CI: 0.6-1.6%) for those interacting with infected individuals in the first 5 days of symptom onset (Cheng et al., 2020), which is similar to the 1.9% attack rate (95% CI 1.8%–2.0%) reported in South Korea (Park et al., 2020).

Current advice treats the larger risk of longer exposure the same, making a 0.13% threshold more conservative because it is calculated to generate a 14-day quarantine for a minimal duration of exposure. However, this is offset by our assuming maximal shedding in calculating this benchmark example. In other words, while this threshold approximates the risk tolerance of current advice, the details of who is recommended for quarantine and for how long will be different in our quantification of total risk than it would be if we were to combine independent binary thresholds for infectious period of transmitters, duration of exposure, and distance to produce a quarantine duration of uniform length. This leads to more consistent treatment of risk to yield a larger benefit in terms of transmission prevented per day of quarantine recommended. Shorter quarantines might significantly reduce the harms imposed by quarantine (Brooks et al., 2020), and increase compliance (Soud et al., 2009, although see McVernon et al., 2011). Quarantining for 14 days post-exposure may be exceptionally challenging for essential workers, individuals without sick leave, or those who would endure significant financial hardship due to lost income.

The assumed fraction of asymptomatic infections affects the discounting of risk. The symptomatic fraction is discounted according to the distribution of incubation periods from exposure to symptom onset, while releasing the asymptomatic fraction from quarantine is not safe until not only onset, but also significant shedding is over (Methods section 2.5.). Our calculations so far assume that 20% of infections are asymptomatic. If we instead assume that 50% infections are asymptomatic, e.g. in a young age group, even a 15-minute contact registered as low attenuation and with peak shedding in the transmitter would require a 16-day quarantine to meet a 0.13% threshold (Fig. 4B). However, if an individual were to test negative during their quarantine, their conditional probability of current or future infectiousness would drop, shortening their quarantine to 13 days for a test with 70% sensitivity (Fig. 4C).

## 4. DISCUSSION

Here we quantify relative risk of infection using experiments to inform the noisy distance-attenuation relationship, and Monte Carlo simulations to inform both this and other sources of variability and uncertainty that affect risk. We roughly calibrate relative infection risk to absolute probability of infection based on limited information from the infection probability of household contacts.

Errors in calibration are likely, but will generally not affect the rank order of risks. For example, adjusting the risk threshold of 0.13% for quarantine will have similar effects to adjusting the value of λ. Knowledge of absolute vs. relative risk does have some effect once some saturation in risk begins to occur, little of which will occur unless much longer durations are recorded.

With 20% cases being asymptomatic and no testing, the risk of current or future infectiousness falls ∼11-fold over the first 14 days of quarantine. Under GAEN v1.5, risk will sometimes differ more between two individuals entering quarantine than when comparing the same individual before vs. after a 14-day quarantine. For this reason, our scheme could recommend quarantines longer than 14 days. Variation in quarantine length is to be expected – if total risk is scored consistently, some quarantines will be longer and others shorter, in order for residual infection probability, conditional on time elapsed without symptoms, to fall below a threshold.

The Covid Watch app is currently programmed either to use a threshold on infection risk to determine 14-day quarantine onset, or on risk of current and future infectiousness to determine both quarantine and duration. Either threshold can be set by public health authorities flexibly in the light of external factors such as level of community transmission, jurisdictional comfort with uncertainty related to digital exposure notifications, and current public health science and recommendations. Communities that have achieved containment might choose to set a stricter threshold, testing individuals once or twice to lower their risk following each negative test. Communities with high prevalence might raise the threshold if it seems likely that the number of quarantine recommendations being issued by the app will cause it to fall out of use, although this issue has not as yet been reported.

When a threshold is set well below the probability that a randomly chosen member of the population is currently infected, it should be recognized that individuals agreeing to download and comply with the recommendations of the app are implicitly agreeing to adhere to higher standards than those implied by the current absence of a general stay-at-home order (Petrie & Masel, 2020). At the time of writing (January 16, 2021), the rate of current infection is ∼5% in Arizona (Gu, 2020). Note that the maximum possible initial infection risk under GAEN version 1 comes to 3.81%. GAEN version 2, by relaxing the 30-minute cap on durations, has made possible resolution among higher risks, although it has complicated approaches to resolve levels of infectiousness.

When the infection risk of the average person in the population is high, we believe that the best solutions are population-level restrictions and closures (Petrie & Masel, 2020). Even when such restrictions are in place, a GAEN app might still have some utility, especially for essential workers. A GAEN app could also be an inferior but still useful option should the political will for population-level restrictions not exist.

As the conditional probability of current or future infectiousness (conditioned on the exposed individual being asymptomatic) falls throughout their quarantine period, messaging can also change. E.g., during the initial high risk days, users might be offered concrete resources such as grocery delivery, or the option to quarantine in a specialized facility in order to protect other household members, before transitioning to self-quarantine once risks falls. Even with self-quarantine, an app might identify the days on which staying home is the highest priority (I.e. days where the potential infectivity may be highest). Messaging considerations are discussed in Supplementary Material Section 3.

We caution that our derived relationship between Bluetooth attenuation and infection risk is extremely approximate and model-dependent. For example, our model calibration assumes that individuals are stationary and that the distance between phones represents the distance between individuals. Additionally, we do not address risk reduction benefits of masks, since mask usage is not captured by the app. Mask filtration efficacies vary by material type, adequacy of fit, and particle size (Pan, Harb, Leng, & Marr, 2020).

Because conservatism vs. permissiveness ultimately depends on the risk threshold, exclusion of mask wearing constitutes overestimation of the relative risk of masked contact, with underestimation of the relative risk of unmasked contact as a corollary. The way to make risk assessment more conservative is to decrease in the risk threshold, while keeping upstream assumptions as accurate as possible. The inability to accurately assess risk with respect to mask use is mitigated by the fact that mask-wearing individuals are likely to have lower risk tolerance, making it more acceptable that a lower implicit risk threshold is applied to them.

Regarding uncertainties in model parameters captured by the app, we have more confidence in our settings of infectiousness levels for symptomatic cases, but very little for asymptomatic cases. These parameters need to be calibrated with real world data on app users who report their app-recorded exposures to manual contact tracing efforts, who then track which users go on to test positive, and who are therefore able to mine the data to quantify the quantitative relationship between exposure details (duration, attenuation, infectiousness) and probability of infection. Transfer of this data to central databases, ideally contact management databases, is critical to improve the targeting of quarantine recommendations to those at highest risk of being infected. Improved risk calibration will make most efficient use of each day of quarantine recommended to reduce transmission.

Short of this, more quantitative data on infectivity would be extremely valuable. Our determination of infectiousness partly relies on the prospective sampling of all individuals in a skilled nursing facility (Arons et al., 2020), where many patients subsequently got sick. Daily samples during similar outbreaks could be used to quantify how shedding varies both among individuals and as a function of time relative to symptom onset. TCID50 data would be ideal, but even Ct values can be valuable for this purpose. However, the fact that the settings we originally chose based infectivity data agreed with later and improved epidemiological approaches is encouraging (Ashcroft et al., 2020; Ferretti, Ledda, et al., 2020).

Without the extended durations provided in version 2 of GAEN, our default calibrations will not recommend quarantine (Fig. 3B) or extended quarantine (Fig. 3C) for contact with less infectious individuals. However, with the duration cap lifted in GAEN version 2, 43 minutes in the ≤50 dB range, 1.43 hours in the 50-60 dB range, or 2.85 hours in the 60-70 dB range with an individual of transmission risk level 2 would be sufficient to trigger quarantine (Figure 3). However, GAEN version 2 provides only two levels of infectiousness, and implementing more as Germany has done (Klingbeil, 2020b, 2020a), requires managing the considerable complexities of separate calculations for shared key servers that need to be interoperable (Justus Benzler, personal communication).

. Limited durations and infectiousness information have been driven by privacy concerns, but this must be weighed against the significant ethical considerations in favor of efficient allocation of quarantine (Singer & Masel, 2020). In Supplementary Section 3, we suggest an alternative method to preserve anonymity, which is to conceal all exposure details from the user’s view. When using variable quarantine duration, this also effectively conceals the date of exposure.

Our framework can be used not only to guide recommendations for who should quarantine and for how long, but also to allocate associated resources including quarantine facilities, grocery delivery and other social support, and priority for access to scarce tests. Both manual contact tracing and digital exposure notification require rapid testing to be effective.

Given limited tests, targeting those at highest risk of infection will do the most good in finding new positive cases who are early enough in the course of infection for these approaches to stem transmission the most.

## Data Availability

The code is available at the github link provided in the text. The Supplementary Datafile is currently available to reviewers and on request, and will be freely available upon journal publication.

## DATA AVAILABILITY

Supplementary Data Table I provides the alpha test data used to calibrate our weights.

## CODE AVAILABILITY

Code and necessary data are accessible under a Creative Commons license at https://github.com/awilson12/risk_scoring

## Supplemental Materials

**Supplementary Table 1.** Parameter values used by app to calculate risk.

| Parameter | Description | Distribution or Point Value |  | References |
| --- | --- | --- | --- | --- |
| $S$ | Viral shedding rate in arbitrary units that are proportional to viral copies/m<sup>3</sup> | 10<sup>1</sup> | 5 days pre- or 8-9 days post-symptom onset | 10-fold range informed by TCID50 measures<sup>1</sup>, timing informed by<sup>1-4</sup> |
|  |  | 10<sup>1.2</sup> | 6-7 days post-symptom onset, or asymptomatic within 2-3 days of test |  |
|  |  | 10<sup>1.4</sup> | 4 days pre- or 5 days post-symptom onset, or asymptomatic within 1 day of test |  |
|  |  | 10<sup>1.6</sup> | 3 days pre- or 4 days post-symptom onset, or asymptomatic on test day |  |
|  |  | 10<sup>1.8</sup> | 2 days pre- or 3 days post-symptom onset |  |
|  |  | 10<sup>2</sup> | 1 day pre- to 2 days post-symptom onset |  |
| $T_{low}, T_{med}, T_{high}$ | Duration of exposure | Durations for Bluetooth attenuations ≤50dB, 50-60dB, 60-70dB, and >70dB are multiplied by weights 2, 1, 0.5, 0 respectively. | | This study |
| $\lambda$ | Probability that one viral particle establishes infection × conversion from arbitrary units | 3.70 x 10<sup>-6</sup> | | Calibrated from secondary attack rate of household contacts = 30%.<sup>5</sup> |
| Fraction of asymptomatic infections | Higher values lead to longer quarantine | 20% for a population, but depends on age |  | <sup>6</sup> |
| Incubation period | Days until symptom onset | Probabilities for {0,1,2...} days = {0,4E-05, 0.011842, 0.088541, 0.181965, 0.207344, 0.174797, 0.123761, 0.081488, 0.051057, 0.031469, 0.018734, 0.011235, 0.006786, 0.00422, 0.002518, 0.001626, 0.000978, 0.000592, 0.000364, 0.000231, 0.00014, 0.000093, 0.000062, 0.00004, 0.000025, 0.000017, 0.000011, 0.000008} |  | <sup>7</sup> |
| Asymptomatic shedding duration | We assume that asymptomatic shedding begins 3 days before what would have been the day of symptom onset if symptomatic, or else immediately upon infection, whichever occurs later, and that shed viral particles are nonviable beyond 12 days | Cumulative probabilities for days {5,6,7...}={0.054054054,0.094594595,0.12162162,0.148648649,0.189189189,0.21621621,0.256756757,1} |  | <sup>8,9</sup> |
| Risk threshold | Set by public health agency, cognizant of limitations in current calibration of $\lambda$ | Benchmark of 0.5% for probability of infection or 0.13% for probability of current or future infectiousness | | This study |

**Supplementary Table 2.** Parameter values used by us to calibrate parameter values in Table 1

| Parameter | Description | Distribution or Point Value | References |
| --- | --- | --- | --- |
| $X$ | Exhalation rate (m<sup>3</sup>/day) | Normal(16.3, 4.15), left-truncated at 9 | <sup>10</sup> |
| $A$ | Cross-sectional area of the mouth. Used to calculate the breath velocity, $U$ from $X$ | Uniform(23, 59) (cm<sup>2</sup>) | <sup>11</sup> |
| $I_y$ | “Lateral intensity” of plume deviation | Uniform(0.08, 0.25) | <sup>12</sup> |
| $I_z$ | “Vertical intensity” of plume deviation | Uniform(0.03, 0.07) | |
| $I$ | Inhalation rate (m<sup>3</sup>/day) | Normal(16.3, 4.15), left-truncated at 9 | <sup>10</sup> |
| $\rho$ | Distance | Sampled from attenuation-distance dataset to inform weights. | This study |
| $\varphi$ | Angle between the z-axis and the xy-plane | Used while informing weights. If $\rho \leq 1$ m, $\varphi = \pi/2$, If $\rho > 1$ m, $\varphi$ randomly sampled from Triangular(min= $\pi/4$, mode= $\pi/2$, max= $3\pi/4$) | 1 m. cutoff for face-to-face interaction informed by <sup>13</sup> |
| $\theta$ | Angle between x and y axes | Used while informing weights. If $\rho \leq 1$ m, $\theta = 0$, If $\rho > 1$ m, $\theta$ randomly sampled from Uniform(0, $2\pi$) | 1 m. cutoff for face-to-face interaction informed by <sup>13</sup> |

**Figure S1.**
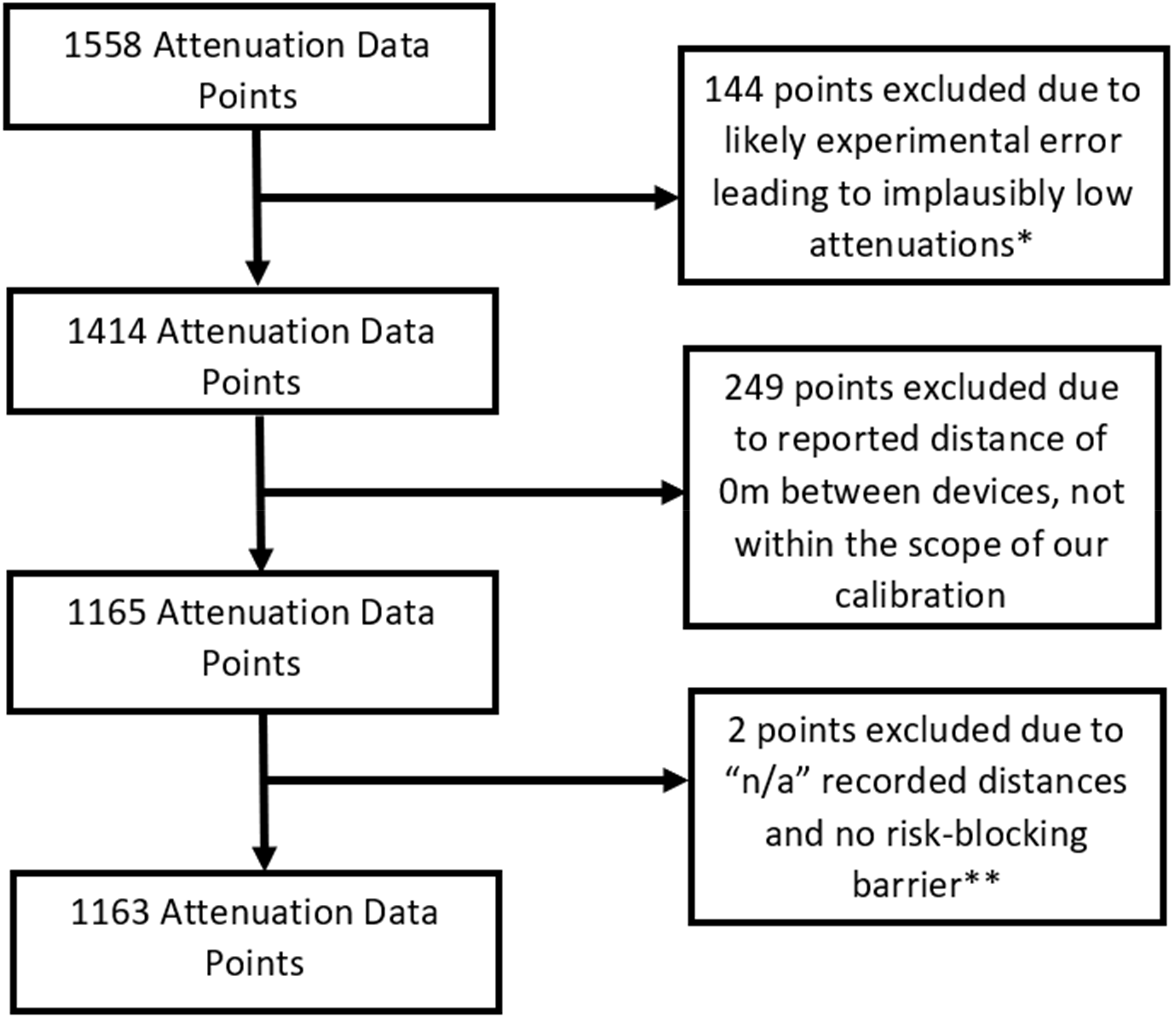
Attenuation data cleaning. *A few attenuation values were implausibly low, always representing one such increment per device per series of attenuation values corresponding to a single test. We believe this is because these testers used the dropdown menu to turn off Bluetooth rather than going to Settings, and this only disables existing Bluetooth connections, causing an anomalously strong signal to be recorded during the period in which the test was being set up. We manually annotated these, totaling 144 datapoints, and excluded them from further analysis, yielding 1414 datapoints. **Distance is not needed for the 128 datapoints taken in the presence of a risk-blocking barrier (e.g. closed car doors or walls), so points marking “N/A” for distance were not excluded for any of these.

### 1.0 Variance in Dose

Our dose response curve is actually the probability of infection as a function of the expectation of dose, rather than dose. To consider the effect of variance in dose, we note that while *E*[1 *-e*^−*λD*^] is analytically intractable with respect to a lognormally distributed dose, there is a viable saddle point approximation described by Rojas-Nandayapa (2008).^14^ While Eq. 5.6 in Rojas-Nandayapa (2008) contains a typo, we use the method given to derive

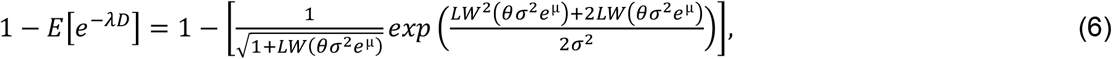

where *LW*(*x*) is the Lambert-W function.

Using this approximation, or the associated importance sampling method^15^ which yields similar results, we can compare the shape for the dose-response curves and consider whether an “effective” *λ* will perform acceptably. Ct counts in Long et al. (2020)^8^ have a standard deviation ∼4, representing an upper bound of a 16-fold difference in viral load, because a difference of 1 Ct in PCR measures represents at most a 2-fold difference in underlying viral load, and because individuals are not sampled at exactly comparable times with respect to the timecourse of shedding. However, variance in dose from causes other than infectiousness, e.g. the intimacy of contact, is not included. With a standard deviation representing a 16-fold difference, we find that by using a value 3/10^th^ of *λ =* 3.70 *E−* 06, we can super-impose the two curves up to 20% infection probability (Figure S2).

**Figure S2.**
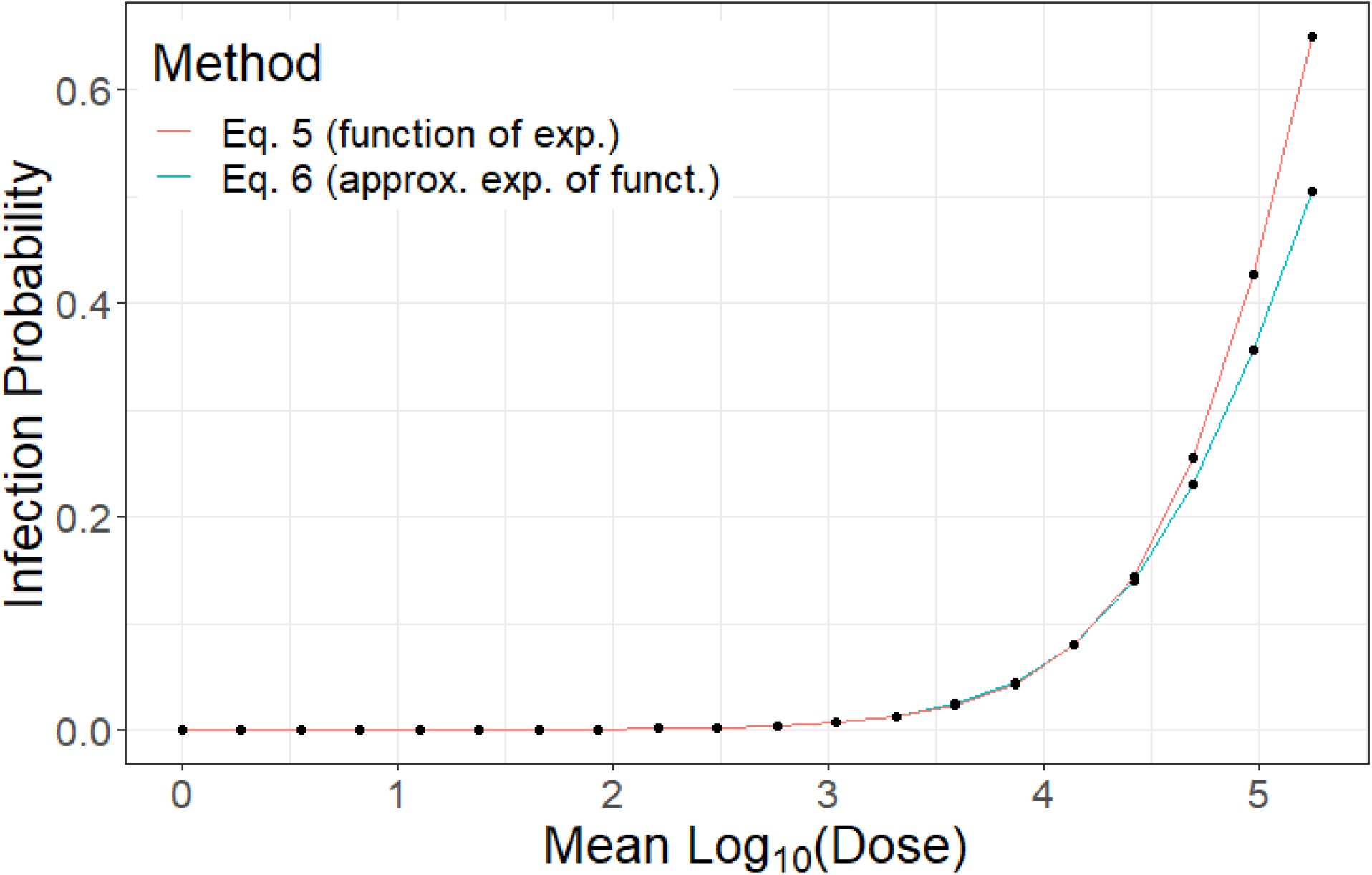
Although our dose-response curve takes the function of an expectation, for low infection probabilities the effect of this is to change the interpretation of the value of ***λ***, which is 3.70 × 10^−6^ when using Eq. 5 but 0.3 times this value when using Eq.6 with a distribution of log-dose with standard deviation corresponding to a 16-fold difference in dose.

### 2.0 Infectiousness

Infectiousness in the GAEN API is a proxy for the magnitude of viral shedding. In both version 1 and version 2 of the Covid Watch app, it is set on the basis of a simple questionnaire administered to users reporting a positive diagnosis. We use this to inform expected shedding *S*. Infectiousness can be encoded by the 8 “Transmission risk levels” in GAEN version 1, and by the two non-zero “infectiousness” levels in GAEN version 2. By repurposing “report type” metadata associated with Temporary Exposure Keys, 8 levels of infectiousness have also been implemented in Germany’s version 2 Corona-Warn-App,^16,17^ requiring separate calculations for shared key servers that need to be interoperable (Justus Benzler, personal communication).

The first question asked by the Covid Watch app to inform infectiousness is, “What day did your symptoms start”? This question has since been made integral to the function of GAEN version 2. A curve fit to known transmission events suggests peak transmission around the day of symptom onset.^18,19^ Estimates of transmission rates as a function of time relative to symptom onset have been used to estimate infectiousness.^20^ However, infectiousness risk as estimated post-symptom onset from transmission rates might be confounded with behavioral changes with symptom onset, leading to the underestimation of post-symptomatic infectiousness.

A second proxy for infectiousness comes from quantitative polymerase chain reaction (PCR). However, this may reflect non-infectious viral remnants, especially late in the course of disease, where the proportion of culture-positive PCR results tends to decrease.^1,2^ We note that this decline is also expected from a simple dose-response curve, where the probability of culture-positivity decreases as the amount of shedding decreases late in infection, i.e. the decline in infectivity might be quantitative rather than qualitative.

Data in which virus was successfully cultured from patient samples is the clearest metric of post-symptom onset infectivity. Arons et al. ^2^ took prospective samples throughout a nursing home, and were able to culture virus from six days before symptom onset until nine days after symptom onset, with little quantitative trend in shedding rate conditional on a positive test. In hospitalized patients, Wölfel et al. ^3^ were unable to isolate live virus from cultures more than 8 days post symptom onset, despite PCR evidence of high shedding. In one case report, live virus has been isolated 18 days after symptom onset, but this seems to be an outlier.^21^ Bullard et al. ^1^ quantified both TCID50 and PCR for 7 days post symptom onset, and saw an approximately 10-fold decline in infectious dose. We note that culture methods may not be sensitive enough to capture low concentrations.^2^ More studies measuring infectivity in a quantitative manner are needed, particularly in populations that represent a broader base of cases of varying ages and health status. Encouragingly, while our infectiousness settings based on culture data did not agree with initial epidemiological estimates of transmission,^22^ they agreed after corrections to the latter were made.^18^

A final source of information comes from detailed Taiwanese contact tracing^23^, who found a 1.0% symptomatic attack rate (95% CI 0.6-1.6%) for those exposed within five days of symptom onset, and 0% (95% CI 0–0.4%) for those exposed after. Risk from exclusively pre-symptomatic exposure was 0.7% (95% CI 0.2%-2.4%). German contact tracing also points to highest transmission risk around the time of symptom onset.^24^

Here we propose the use of 6 infectiousness levels in the GAEN API, evenly spaced on a log scale between 10 and 100 in arbitrary units, reserving the use of 2 levels for future functionality e.g. regarding superspreaders. Infectiousness levels could also be manipulated for testing purposes, e.g. to help learn, if individuals voluntarily share exposure details centrally, how infectiousness varies in the real world as a function of symptomatic status and timing. We note that most GAEN version apps launched with only one level of infectiousness, and that GAEN version 2 is designed in anticipation of the use of only two the use of only two. While Germany has succeeded in repurposing metadata to provide more, this increases the complexity of interoperability. Given that a systematic 10-fold difference in TCID50 has been observed, supported by epidemiological data, more than 2 levels seem warranted, and making them an intrinsic part of the GAEN system would increase the likelihood of their being used.

Based on a holistic reading of the four sources of evidence described above, we assign the maximum level of 6 from one day pre-symptom onset to two days post-symptom onset. Five days before symptom onset we assign level 1, four days before we assign level 3, three days before level 4, and two days before level 5. Three days after symptom onset we assign level 5, four days after level 4, five days after level 3, 6-7 days after level 2, and 8-9 days after level 1. Our termination at 9 days is based on current CDC guidance.^9^ Pre-symptomatic infectiousness assignments can be further refined in cases when the date of exposure is known, given that the infectious period is longer for longer incubation periods.^19^

For users who report a positive test but no symptoms, there is likely a reason they were tested, and so we ask for the most likely day of exposure, if known. If provided, we assume that shedding did not begin until two days after exposure, at the earliest. We also ask for the date of sampling for the positive test (which can be reported by the healthcare provider rather than the app user) and assume peak shedding at around this time. Subject to the constraint from day of exposure, we assign infectiousness 4 to the day of the test, 3 to the day before and after, and level 2 to dates between 2 and 3 days of the test. There is some evidence that viral shedding is lower in asymptomatic vs. symptomatic cases,^25^ while another study indicates the shedding magnitudes may be similar.^26^ Note that we assume that those with no symptoms at the time they receive a positive test result are asymptomatic rather than pre-symptomatic. We recommend allowing app users to report symptom onset after the fact and trigger a change to previously reported infectiousness. GAEN version 2 TEK revocation functionality makes this possible.

### 3.0 Considerations in recommending and messaging variable quarantine durations

The need for consistent guidance to the public is an important consideration for implementing tailored risk scoring and modified quarantine recommendations. If for the sake of a consistency, a public health authority is not willing to authorize variable quarantine recommendations, as was the case in Arizona when this work was conducted, but only 0 or 14 day quarantines from time of the last individually significant exposure, then the threshold for going into quarantine at all would need to become more strict in order to maintain the same overall risk among the population under quarantine. In other words, retaining the same average probability of current or future infectiousness among the quarantined population would require some exposed individuals to no longer go into quarantine at all, in addition to others lengthening their quarantine out to 14 days. With a binary 0 or 14 day quarantine, the amount by which disease transmission is prevented per day of quarantine will be lower. Considerations are similar for alternative CDC guidelines^27^ for 10 day quarantine or 7 day given a negative test in the last 48 hours.

Alternatively, to avoid mixed messaging regarding quarantine even while the app recommends quarantine of variable duration, one option is to suppress all details about individual exposures from the user’s view, including their date. This has the additional advantage of decreasing the risk that users will be able to guess who exposed them, further preserving privacy.

The app can communicate the risk of infectiousness either as a simple recommendation for which days to quarantine, or also as a quantitative score in order to “game-ify” the process of quarantine and give users positive feedback for each day they succeed in remaining at home until risk falls to a lower level. Further research is needed to assess the most effective messaging strategies. E.g., the app could display both current and projected risk of infectiousness on a simple scale of 1 to 10, so users can see how that risk will fall with each day of quarantine. This visualization might change perceptions. E.g., an individual who wants to comply with a 14-day quarantine, but does not feel able to, might rush out to get groceries before starting their quarantine in earnest, while shedding virus pre-symptomatically. Visualizing projected risk into the future would then give the message that if the exposed individual can only make do for one more day before leaving the home for essentials, that will help, because if they do not develop symptoms, their risk will be lower even after a single day longer. Risk communication in an app could focus on day to day coaxing of this form. Basing quarantine recommendation on a threshold for the expected number of onward transmissions per day is more socially optimal than a threshold on the conditional probability of infectiousness as described here.^28^

Conflicting messages can still arise if manual contact tracers trace an individual who also received an exposure notification. In this case, it is likely that the two recommend different end dates for quarantine. While this is to be expected from our procedure for recommending variable quarantine durations, we note that even if the app were to issue 14 day quarantine recommendations only, it could still arise because the individual has been exposed more than once, on different days, and the manual contact tracer is following up an infected individual who may not have used the app. Until there is reliable data on app performance, we recommend that the manual contact tracer’s protocol should override whatever the app says. Should the app turn out to perform well, an alternative procedure might eventually be to go with whichever protocol recommends the longer quarantine. An intermediate possibility is for the manual contact tracer to ask for exposure notification details, to determine whether it may be a different exposure to the one being manually traced. There may also be conflicts in protocols for the timing of testing.

Note that with symptom onset sometimes as early as two days after exposure, and given the possibility of pre-symptomatic shedding, and the possibility of confusion regarding who infected whom, we currently ignore the possibility that shedding might not yet have begun. Current testing turnaround times are mostly long, making this reasonable. However, if same-day tests become more widely available, our approach could be extended to directly communicate the risk of current infectiousness, whose calculation is described in Petrie & Masel (manuscript in prep.)^29^, rather than as is currently the case, the risk of current or future infectiousness. A significantly lower risk of infectiousness will be present on the day of exposure and perhaps also the day after. Delays in going into public to prepare for a long quarantine could inadvertently lead to pushing individuals past the latent period before they go into public; displaying a full projected timeline of the projected risk of infectiousness could avert this, at the risk of significantly more complex messaging than “stay home until Friday”. The use of current infectiousness would prevent the app from occasionally recommending quarantines of less than 5 days when initial risk is already very near the threshold.

## REFERENCES

Ai, T., Yang, Z., Hou, H., Zhan, C., Chen, C., Lv, W., … Xia, L. (2020). Correlation of Chest CT and RT-PCR Testing in Coronavirus Disease 2019 (COVID-19) in China: A Report of 1014 Cases. Radiology, 296(2), E32–E40. https://doi.org/10.1148/radiol.2020200642

Aleta, A., Martin-Corral, D., Pastore Y Piontti, A., Ajelli, M., Litvinova, M., Chinazzi, M., … Moreno, Y. (2020). Modeling the impact of social distancing, testing, contact tracing and household quarantine on second-wave scenarios of the COVID-19 epidemic. MedRxiv. https://doi.org/10.1101/2020.05.06.20092841

Armbruster, B., & Brandeau, M. L. (2007). Contact tracing to control infectious disease: When enough is enough. Health Care Management Science, 10(4), 341–355. https://doi.org/10.1007/s10729-007-9027-6

Arons, M. M., Hatfield, K. M., Reddy, S. C., Kimball, A., James, A., Jacobs, J. R., … Jernigan, J. A. (2020). Presymptomatic SARS-CoV-2 infections and transmission in a skilled nursing facility. The New England Journal of Medicine, 382(22), 2081–2090. https://doi.org/10.1056/NEJMoa2008457

Ashcroft, P., Huisman, J. S., Lehtinen, S., Bouman, J. A., Althaus, C. L., Regoes, R. R., & Bonhoeffer, S. (2020). COVID-19 infectivity profile correction. Swiss Medical Weekly, (150), w20336. https://doi.org/10.4414/smw.2020.20336

Bi, Q., Wu, Y., Mei, S., Ye, C., Zou, X., Zhang, Z., … Feng, T. (2020). Epidemiology and transmission of COVID-19 in 391 cases and 1286 of their close contacts in Shenzhen, China: a retrospective cohort study. The Lancet Infectious Diseases, 20(8), 911–919. https://doi.org/10.1016/S1473-3099(20)30287-5

Brooks, S. K., Webster, R. K., Smith, L. E., Woodland, L., Wessely, S., Greenberg, N., & Rubin, G. J. (2020). The psychological impact of quarantine and how to reduce it: rapid review of the evidence. The Lancet, 395(10227), 912–920. https://doi.org/10.1016/S0140-6736(20)30460-8

Brusca, S., Famoso, F., Lanzafame, R., Mauro, S., Garrano, A. M. C., & Monforte, P. (2016). Theoretical and experimental study of Gaussian plume model in small scale system. Energy Procedia, 101, 58–65. https://doi.org/10.1016/j.egypro.2016.11.008

Buitrago-Garcia, D. C., Egli-Gany, D., Counotte, M. J., Hossmann, S., Imeri, H., Salanti, G., & Low, N. (2020). Asymptomatic SARS-CoV-2 infections: a living systematic review and meta-analysis, version 3. MedRxiv. https://doi.org/10.1101/2020.04.25.20079103

Bullard, J., Dust, K., Funk, D., Strong, J. E., Alexander, D., Garnett, L., … Poliquin, G. (2020). Predicting infectious SARS-CoV-2 from diagnostic samples. Clinical Infectious Diseases. https://doi.org/10.1093/cid/ciaa638

Centers for Disease Control and Prevention. (2020). Coronavirus Disease 2019 Community-Related Exposures. Retrieved January 7, 2020, from https://www.cdc.gov/coronavirus/2019-ncov/php/public-health-recommendations.html

Chau, N. V. V., Thanh Lam, V., Thanh Dung, N., Yen, L. M., Minh, N. N. Q., Hung, L. M., … Tan L. Van. (2020). The natural history and transmission potential of asymptomatic SARS-CoV-2 infection. Clinical Infectious Diseases, 71(10), 2679–2687. https://doi.org/10.1093/cid/ciaa711

Chen, W., Zhang, N., Wei, J., Yen, H. L., & Li, Y. (2020). Short-range airborne route dominates exposure of respiratory infection during close contact. Building and Environment, 176, 106859. https://doi.org/10.1016/j.buildenv.2020.106859

Chen, X., Zhang, Y., Zhu, B., Zeng, J., Hong, W., He, X., … Wang, J. (2020). Associations of Clinical Characteristics and Antiviral Drugs with Viral RNA Clearance in Patients with COVID-19 in Guangzhou, China. MedRxiv. https://doi.org/10.1101/2020.04.09.20058941

Cheng, H. Y., Jian, S. W., Liu, D. P., Ng, T. C., Huang, W. T., & Lin, H. H. (2020). Contact tracing assessment of COVID-19 transmission dynamics in Taiwan and risk at different exposure periods before and after symptom onset. JAMA Internal Medicine, 180(9), 1156– 1163. https://doi.org/10.1001/jamainternmed.2020.2020

Chu, D. K., Akl, E. A., Duda, S., Solo, K., Yaacoub, S., Schünemann, H. J., … Reinap, M. (2020). Physical distancing, face masks, and eye protection to prevent person-to-person transmission of SARS-CoV-2 and COVID-19: a systematic review and meta-analysis. The Lancet, 395(10242), 1973–1987. https://doi.org/10.1016/S0140-6736(20)31142-9

Curmei, M., Ilyas, A., Evans, O., & Steinhardt, J. (2020). Estimating household transmission of SARS-CoV-2. MedRxiv. https://doi.org/10.1101/2020.05.23.20111559

Davies, N. G., Klepac, P., Liu, Y., Prem, K., Jit, M., & Eggo, R. M. (2020). Age-dependent effects in the transmission and control of COVID-19 epidemics. Nature Medicine, 26(8), 1205–1211. https://doi.org/10.1038/s41591-020-0962-9

European Centre for Disease Prevention and Control. (2020). Contact tracing: public health management of persons, including healthcare workers, who have had contact with COVID-19 cases in the European Union – third update. Retrieved January 7, 2020, from https://www.ecdc.europa.eu/en/covid-19-contact-tracing-public-health-management

Farrell, S., & Leith, D. J. (2020). Pairwise Handset Types and Orientations Are Sufficient to Blur Exposure Notification Thresholds. Retrieved August 6, 2020, from Trinity College Dublin, Ireland website: https://down.dsg.cs.tcd.ie/tact/posorient.pdf

Feng, Y., Marchal, T., Sperry, T., & Yi, H. (2020). Influence of wind and relative humidity on the social distancing effectiveness to prevent COVID-19 airborne transmission : A numerical study. Journal of Aerosol Science, 147, 105585. https://doi.org/10.1016/j.jaerosci.2020.105585

Fennelly, K. P. (2020). Particle sizes of infectious aerosols: implications for infection control. The Lancet Respiratory Medicine, 8(9), 914–924. https://doi.org/10.1016/S2213-2600(20)30323-4

Ferretti, L., Ledda, A., Wymant, C., Zhao, L., Ledda, V., Abeler-, L., … Fraser, C. (2020). The timing of COVID-19 transmission. MedRxiv. https://doi.org/10.1101/2020.09.04.20188516

Ferretti, L., Wymant, C., Kendall, M., Zhao, L., Nurtay, A., Abeler-Dörner, L., … Fraser, C. (2020). Quantifying SARS-CoV-2 transmission suggests epidemic control with digital contact tracing. Science, 368(6491), eabb6936. https://doi.org/10.1126/science.abb6936

Fetzer, T., & Graeber, T. (2020). Does contact tracing work? Quasi-experimental evidence from an Excel error in England. MedRxiv. https://doi.org/10.1101/2020.12.10.20247080

Fraser, C., Abeler-Dörner, L., Ferretti, L., Parker, M., Kendall, M., & Bonsall, D. (2020). Digital contact tracing: comparing the capabilities of centralised and decentralised data architectures to effectively suppress the COVID-19 epidemic whilst maximising freedom of movement and maintaining privacy. Introduction 2. Retrieved September 5, 2020, from https://github.com/BDI-pathogens/covid19-instant_tracing/blob/master/Centralised%20and%20decentralised%20systems%20for%20contact%20tracing.pdf.

Gu, Y. (2020). covid19-projections.com. Retrieved September 5, 2020, from https://youyanggu.com/about

Guo, S., Yu, J., Shi, X., Wang, H., Xie, F., Gao, X., & Jiang, M. (2019). Droplet-transmitted infection risk ranking based on close proximity interaction. Frontiers in Neurorobotics, 13(113). https://doi.org/10.3389/fnbot.2019.00113

Haas, C. N., Rose, J. B., & Gerba, C. P. (1999). Quantitative Microbial Risk Assessment. John Wiley and Sons.

Hamner, L., Dubbel, P., Capron, I., Ross, A., Jordan, A., Lee, J., … Leibrand, H. (2020). High SARS-CoV-2 attack rate following exposure at a choir practice - Skagit County, Washington, March 2020. Morbidity and Mortality Weekly Report High, 69(19), 606–610. https://doi.org/10.15585/mmwr.mm6919e6

Hellewell, J., Russell, T. W., The SAFER Investigators and Field Study Team, The Crick COVID-19 Consortium, CMMID COVID-19 working group, Beale, R., … Kucharski, A. J. (2020). Estimating the effectiveness of routine asymptomatic PCR testing at different frequencies for the detection of SARS-CoV-2 infections. MedRxiv. https://doi.org/10.1101/2020.11.24.20229948

Hu, Z., Song, C., Xu, C., Jin, G., Chen, Y., Xu, X., … Shen, H. (2020). Clinical characteristics of 24 asymptomatic infections with COVID-19 screened among close contacts in Nanjing, China. Science China Life Sciences, 63(5), 706–711. https://doi.org/10.1007/s11427-020-1661-4

Jain, S., Islam, T., Chowdhury, B., Chen, Y., & Son, Y. (2021). Agent-based simulation to evaluate various entrance and exit policies and contact-caused risks in classrooms under a pandemic situation. In preparation.

Jang, S., Han, S. H., & Rhee, J.-Y. (2020). Cluster of coronavirus disease associated with fitness dance classes, South Korea. Emerging Infectious Diseases, 26(8), 1917–1920. https://doi.org/10.3201/eid2608.200633

Jones, N. R., Qureshi, Z. U., Temple, R. J., Larwood, J. P. J., Greenhalgh, T., & Bourouiba, L. (2020). Two metres or one: what is the evidence for physical distancing in covid-19? BMJ, 370, m3223. https://doi.org/10.1136/bmj.m3223

Kendall, M., Milsom, L., Abeler-dörner, L., Wymant, C., Ferretti, L., Briers, M., … Fraser, C. (2020). Epidemiological changes on the Isle of Wight after the launch of the NHS Test and Trace programme: a preliminary analysis. The Lancet Digital Health, 2(12), E658–E666. https://doi.org/10.1016/S2589-7500(20)30241-7

Klingbeil, T. (2020a). corona-warn-app / cwa-documentation / images / risk_calculation / client_interpretation.pdf. Retrieved January 6, 2021, from GitHub website: https://github.com/corona-warn-app/cwa-documentation/blob/master/images/risk_calculation/client_interpretation.pdf

Klingbeil, T. (2020b). corona-warn-app / cwa-documentation / images / risk_calculation / server_encoding.pdf. Retrieved January 6, 2021, from GitHub website: https://github.com/corona-warn-app/cwa-documentation/blob/master/images/risk_calculation/server_encoding.pdf

Kretzschmar, M. E., Rozhnova, G., Bootsma, M. C. J., van Boven, M., van de Wijgert, J. H. H. M., & Bonten, M. J. M. (2020). Impact of delays on effectiveness of contact tracing strategies for COVID-19: a modelling study. The Lancet. Public Health, 5(8), e452–e459. https://doi.org/10.1016/S2468-2667(20)30157-2

Kucharski, A. J., Klepac, P., Conlan, A., Kissler, S. M., Tang, M., Fry, H., … Edmunds, J. (2020). Effectiveness of isolation, testing, contact tracing and physical distancing on reducing transmission of SARS-CoV-2 in different settings: a mathematical modelling study. The Lancet. Infectious Diseases, (10), 1151–1160. https://doi.org/10.1101/2020.04.23.20077024

Kucirka, L. M., Lauer, S. A., Laeyendecker, O., Boon, D., & Lessler, J. (2020). Variation in false-negative rate of reverse transcriptase polymerase chain reaction–Based SARS-CoV-2 tests by time since exposure. Annals of Internal Medicine. https://doi.org/10.7326/m20-1495

Lauer, S. A., Grantz, K. H., Bi, Q., Jones, F. K., Zheng, Q., Meredith, H. R., … Lessler, J. (2020). The incubation period of coronavirus disease 2019 (CoVID-19) from publicly reported confirmed cases: Estimation and application. Annals of Internal Medicine, 172(9), 577–582. https://doi.org/10.7326/M20-0504

Leckie, J. O., Naylor, K. A., Canales, R. A., Ferguson, A. C., Cabrera, N. L., Hurtado, A. L., … Vieira, V. M. (2000). Quantifying Children’s Microlevel Activity Data from Existing Videotapes” by Exposure Research Group at Stanford University for the U.S. Environmental Protection Agency. Report No: U2F112OT-RT-99-001182. Retrieved from https://cfpub.epa.gov/ols/catalog/advanced_full_record.cfm?&FIELD1=AUTHOR&INPUT1=FERGUSONANDC.ANDR.&TYPE1=ALL&LOGIC1=AND&COLL=&SORT_TYPE=MTIC&item_count=5

Lednicky, J. A., Lauzardo, M., Fan, Z. H., Jutla, A., Tilly, T. B., Gangwar, M., … Wu, C. (2020). Viable SARS-CoV-2 in the air of a hospital room with COVID-19 patients. International Journal of Infectious Diseases, 100, 476–482. https://doi.org/10.1101/2020.08.03.20167395

Long, Q., Tang, X., Shi, Q., Li, Q., Deng, H., Yuan, J., … Huang, A. (2020). Clinical and immunological assessment of asymptomatic SARS-CoV-2 infections. Nature Medicine, 26(8), 1200–1204. https://doi.org/10.1038/s41591-020-0965-6

Lythgoe, K. A., Hall, M., Ferretti, L., de Casare, M., MacIntyre-Cockett, G., Trebes, A., … Golubchik, T. (2020). Within-host genomics of SARS-CoV-2. BioRxiv. https://doi.org/10.1101/2020.05.28.118992

Ma, J., Qi, X., Chen, H., Li, X., Zhan, Z., Wang, H., … Yao, M. (2020). Exhaled breath is asignificant source of SARS-CoV-2 emission. MedRxiv, https://doi.org/10.1101/2020.05.31.20115154

McVernon, J., Mason, K., Petrony, S., Nathan, P., LaMontagne, A. D., Bentley, R., … Kavanagh, A. (2011). Recommendations for and compliance with social restrictions during implementation of school closures in the early phase of the influenza A (H1N1) 2009 outbreak in Melbourne, Australia. BMC Infectious Diseases, 11. https://doi.org/10.1186/1471-2334-11-257

Ottino-Loffler, B., Scott, J. G., & Strogatz, S. H. (2017). Evolutionary dynamics of incubation periods. ELife, 6, e30212. https://doi.org/10.7554/eLife.30212

Pan, J., Harb, C., Leng, W., & Marr, L. C. (2020). Inward and outward effectiveness of cloth masks, a surgical mask, and a face shield. MedRxiv. https://doi.org/10.1101/2020.11.18.20233353

Park, Y. J., Choe, Y. J., Park, O., Park, S. Y., Kim, Y.-M., Kim, J., … Jeong, E. K.; COVID-19 National Emergency Response Center, Epidemiology and Case Management Team. (2020). Contact tracing during coronavirus disease outbreak, South Korea, 2020. Emerging Infectious Disease Journal, 26(10), 2465–2468. https://doi.org/10.3201/eid2610.201315

Petrie J, Masel J. The economic value of quarantine is higher at lower case prevalence, with quarantine justified at lower risk of infection. medRxiv 2020

Petrie, J., & Masel, J. (2021). Quarantine Optimization. In preparation.

Petrie, J., Nurtay, A., Ferretti, L., Fraser, C., & Masel, J. (2021). Optimal Testing Timing During Quarantine Depends on How Compliance Changes with Test Results. In preparation.

Prather, K. A., Wang, C. C., & Schooley, R. T. (2020). Reducing transmission of SARS-CoV-2. Science, 368(6498), 1422–1424.

Qureshi, Z., Jones, N., Temple, R., Larwood, J. P., Greenhalgh, T., & Bourouiba, L. (2020). What is the evidence to support the 2-metre social distancing rule to reduce COVID-19 transmission? Retrieved July 15, 2020, from CEBM website: https://www.cebm.net/covid-19/what-is-the-evidence-to-support-the-2-metre-social-distancing-rule-to-reduce-covid-19-transmission/

Rea, E., Laflèche, J., Stalker, S., Guarda, B. K., Shapiro, H., Johnson, I., … Eliasziw, M. (2007). Duration and distance of exposure are important predictors of transmission among community contacts of Ontario SARS cases. Epidemiology and Infection, 135(6), 914–921. https://doi.org/10.1017/S0950268806007771

Salathé, M., Althaus, C. L., Anderegg, N., Antonioli, D., Ballouz, T., Bugnion, E., … von Wyl, V. (2020). Early evidence of effectiveness of digital contact tracing for SARS-CoV-2 in Switzerland. Swiss Medical Weekly. https://doi.org/10.4414/smw.2020.20457

Salathé, M., Kazandjieva, M., Lee, J. W., Levis, P., Feldman, M. W., & Jones, J. H. (2010). A high-resolution human contact network for infectious disease transmission. Proceedings of the National Academy of Sciences of the United States of America, 107(51), 22020–22025. https://doi.org/10.1073/pnas.1009094108

Setti, L., Passarini, F., De Gennaro, G., Barbieri, P., Perrone, M. G., Borelli, M., … Miani, A. (2020). Airborne transmission route of covid-19: Why 2 meters/6 feet of inter-personal distance could not be enough. International Journal of Environmental Research and Public Health, 17(8), e2932. https://doi.org/10.3390/ijerph17082932

Singer, P., & Masel, J. (2020, September 3). How (Not) to Fight COVID 19. Project Syndicate. Retrieved from https://www.project-syndicate.org/commentary/tech-solutions-to-targeting-covid19-quarantine-by-peter-singer-and-joanna-masel-2020-08

Soud, F. A., Cortese, M. M., Curns, A. T., Edelson, P. J., Bitsko, R. H., Jordan, H. T., … Dayan, G. H. (2009). Isolation compliance among university students during a mumps outbreak, Kansas 2006. Epidemiology and Infection, 137(1), 30–37. https://doi.org/10.1017/S0950268808000629

U.S. Environmental Protection Agency. (2011). Exposure Factors Handbook 2011 Edition (EPA/600/R-09/052F). Retrieved from https://cfpub.epa.gov/ncea/risk/recordisplay.cfm?deid=236252

Von Arx, S., Becker-Mayer, I., Blank, D., Colligan, J., Fenwick, R., Hittle, M., … Xue, H. (2020). Slowing the Spread of Infectious Diseases Using Crowdsourced Data. Retrieved August 19, 2020, from Covid Watch website: https://blog.covidwatch.org/en/covid-watch-whitepaper-using-crowdsourced-data-to-slow-virus-spread

Wanger, J. (2020). Configuring Exposure Notification Risk Scores. Retrieved January 10, 2021, from GitHub website: https://github.com/lfph/gaen-risk-scoring/blob/main/risk-scoring.md

Webster, R. K., Brooks, S. K., Smith, L. E., Woodland, L., Wessely, S., & Rubin, G. J. (2020). How to improve adherence with quarantine: rapid review of the evidence. Public Health, 182, 163–169. https://doi.org/10.1016/j.puhe.2020.03.007

Wei, Y., Wei, L., Liu, Y., Huang, L., Shen, S., Zhang, R., … Chen, F. (2020). A systematic review and meta-analysis reveals long and dispersive incubation period of COVID-19. MedRxiv. https://doi.org/10.1101/2020.06.20.20134387

Western Engineering. (n.d.). Self-study notes - GAUSSIAN PLUMES. Retrieved June 7, 2020, from https://www.eng.uwo.ca/people/esavory/Gaussian plumes.pdf

World Health Organization. (2020). Considerations for quarantine of individuals in the context of containment for coronavirus disease (COVID-19). Retrieved July 1, 2020, from https://www.who.int/publications/i/item/considerations-for-quarantine-of-individuals-in-the-context-of-containment-for-coronavirus-disease-(covid-19)

Xiao, T., Wang, Y., Yuan, J., Ye, H., Wei, L., Wang, H., … Zhang, Z. (2020). Early viral clearance and antibody kinetics of COVID-19 among asymptomatic carriers. MedRxiv. https://doi.org/10.1101/2020.04.28.20083139

Yang, R., Gui, X., & Xiong, Y. (2020). Comparison of clinical characteristics of patients with asymptomatic vs symptomatic coronavirus disease 2019 in Wuhan, China. JAMA Network Open, 3(5), e2010182. https://doi.org/10.1001/jamanetworkopen.2020.10182

Yang, W., Elankumaran, S., & Marr, L. C. (2011). Concentrations and size distributions of airborne influenza A viruses measured indoors at a health centre, a day-care centre and on aeroplanes. Journal of the Royal Society, Interface, 8(61), 1176–1184. https://doi.org/10.1098/rsif.2010.0686

Yang, Y., Yang, M., Shen, C., Wang, F., Yuan, J., Li, J., … Liu, Y. (2020). Evaluating the accuracy of different respiratory specimens in the laboratory diagnosis and monitoring the viral shedding of 2019-nCoV infections. MedRxiv, 2020.02.11.20021493. https://doi.org/10.1101/2020.02.11.20021493

Zhang, N., Su, B., Chan, P. T., Miao, T., Wang, P., & Li, Y. (2020). Infection spread and high-resolution detection of close contact behaviors. International Journal of Environmental Research and Public Health, 17(4), 1445. https://doi.org/10.3390/ijerph17041445

## References

1 Bullard J, Dust K, Funk D, et al. Predicting infectious SARS-CoV-2 from diagnostic samples. Clin Infect Dis 2020; 71:2663–2666. DOI:10.1093/cid/ciaa638.

2 Arons MM, Hatfield KM, Reddy SC, et al. Presymptomatic SARS-CoV-2 infections and transmission in a skilled nursing facility. N Engl J Med 2020; 382: 2081–90.

3 Wölfel R, Corman VM, Guggemos W, et al. Virological assessment of hospitalized patients with COVID-2019. Nature 2020; 581: 465–9.

4 Ashcroft P, Huisman JS, Lehtinen S, et al. COVID-19 infectivity profile correction. Swiss Med Wkly 2020; 150: w20336.

5 Curmei M, Ilyas A, Evans O, Steinhardt J. Estimating household transmission of SARS-CoV-2. medRxiv 2020. DOI:10.1101/2020.05.23.20111559.

6 Buitrago-Garcia DC, Egli-Gany D, Counotte MJ, et al. Asymptomatic SARS-CoV-2 infections: a living systematic review and meta-analysis, version 3. medRxiv 2020. DOI:10.1101/2020.04.25.20079103.

7 Lauer SA, Grantz KH, Bi Q, et al. The incubation period of coronavirus disease 2019 (CoVID-19) from publicly reported confirmed cases: Estimation and application. Ann Intern Med 2020; 172: 577–82.

8 Long Q, Tang X, Shi Q, et al. Clinical and immunological assessment of asymptomatic SARS-CoV-2 infections. Nat Med 2020; 26: 1200–4.

9 Centers for Disease Control and Prevention. Duration of Isolation & Precautions for Adults. 2020. https://www.cdc.gov/coronavirus/2019-ncov/hcp/duration-isolation.html#cecommendations (accessed July 27, 2020).

10 U.S. Environmental Protection Agency. Exposure Factors Handbook 2011 Edition (EPA/600/R-09/052F). Washington, DC, 2011 https://cfpub.epa.gov/ncea/risk/recordisplay.cfm?deid=236252.

11 Leckie JO, Naylor KA, Canales RA, et al. Quantifying Children’s Microlevel Activity Data from Existing Videotapes” by Exposure Research Group at Stanford University for the U.S. Environmental Protection Agency. 2000 https://cfpub.epa.gov/ols/catalog/advanced_full_record.cfm?&FIELD1=AUTHOR&INPUT1=FERGUSONANDC.ANDR.&TYPE1=ALL&LOGIC1=AND&COLL=&SORT_TYPE=MTIC&item_count=5.

12 Western Engineering. Self-study notes - GAUSSIAN PLUMES. https://www.eng.uwo.ca/people/esavory/Gaussian plumes.pdf (accessed June 7, 2020).

13 Zhang N, Su B, Chan PT, Miao T, Wang P, Li Y. Infection spread and high-resolution detection of close contact behaviors. Int J Environ Res Public Health 2020; 17: 1445.

14 Rojas-Nandayapa L. Risk Probabilities: Asymptotics and Simulation. 2008. Aarhus University. Retrieved from https://pure.au.dk/portal/en/publications/risk-probabilities(8d90e9a0-db1a-11dd-9710-000ea68e967b)/export.html.

15 Asmussen S, Jensen JL, Rojas-Nandayapa L. On the Laplace Transform of the Lognormal Distribution. Methodol Comput Appl Probab 2016; 18: 441–58.

16 Klingbeil T. corona-warn-app / cwa-documentation / images / risk_calculation / server_encoding.pdf. GitHub. 2020. https://github.com/corona-warn-app/cwa-documentation/blob/master/images/risk_calculation/server_encoding.pdf (accessed Jan 6, 2021).

17 Klingbeil T. bcorona-warn-app / cwa-documentation/images/risk_calculation/client_interpretation.pdf. GitHub. 2020. https://github.com/corona-warn-app/cwa-documentation/blob/master/images/risk_calculation/client_interpretation.pdf (accessed Jan 6, 2021).

18 He X, Lau EHY, Wu P, et al. Author Correction: Temporal dynamics in viral shedding and transmissibility of COVID-19. Nat Med 2020; In print.

19 Ferretti L, Ledda A, Wymant C, et al. The timing of COVID-19 transmission. medRxiv 2020. DOI:10.1101/2020.09.04.20188516.

20 CWA Team. corona-warn-app/cwa-documentation/transmission_risk.pdf. GitHub. 2020. https://github.com/corona-warn-app/cwa-documentation/blob/master/transmission_risk.pdf (accessed Jan 6, 2021).

21 Liu W Da, Chang SY, Wang JT, et al. Prolonged virus shedding even after seroconversion in a patient with COVID-19. J Infect 2020. 81: 318–356. DOI:10.1016/j.jinf.2020.03.063.

22 He X, Lau EHY, Wu P, et al. Temporal dynamics in viral shedding and transmissibility of COVID-19. Nat Med 2020; 26: 672–5.

23 Cheng HY, Jian SW, Liu DP, Ng TC, Huang WT, Lin HH. Contact tracing assessment of COVID-19 transmission dynamics in Taiwan and risk at different exposure periods before and after symptom onset. JAMA Intern Med 2020; 180: 1156–63.

24 Böhmer MM, Buchholz U, Corman VM, et al. Investigation of a COVID-19 outbreak in Germany resulting from a single travel-associated primary case: a case series. Lancet Infect Dis 2020; 20: 920–928. DOI:10.1016/S1473-3099(20)30314-5.

25 McDonald J. Unpacking WHO’s Asymptomatic COVID-19 Transmission Comments. FactCheck.org. 2020. https://www.factcheck.org/2020/06/unpacking-whos-asymptomatic-covid-19-transmission-comments/ (accessed June 16, 2020).

26 Lavezzo E, Franchin E, Ciavarella C, et al. Suppression of a SARS-CoV-2 outbreak in the Italian municipality of Vo’. Nature 2020; 584: 425–429. DOI:10.1038/s41586-020-2488-1.

27 Centers for Disease Control and Prevention. Options to Reduce Quarantine for Contacts of Persons with SARS-CoV-2 Infection Using Symptom Monitoring and Diagnostic Testing. 2020. https://www.cdc.gov/coronavirus/2019-ncov/more/scientific-brief-options-to-reduce-quarantine.html (accessed Jan 10, 2020).

28 Petrie J, Masel J. The economic value of quarantine is higher at lower case prevalence, with quarantine justified at lower risk of infection. medRxiv 2020. DOI:10.1101/2020.11.24.20238204.

29 Petrie J, Masel J. Quarantine Optimization. 2021; In preparation.

